# Causally-anchored multi-omic deep learning recovers exercise-responsive and ageing-causal genes from human physical activity

**DOI:** 10.64898/2025.12.26.25343061

**Authors:** Ciara G. Juan, Lazaros Ntasis

## Abstract

Physical activity is among the most robust epidemiological correlates of reduced mortality and multi-morbidity, yet the molecular mechanisms through which exercise exerts these effects in humans remain incompletely resolved. This study combines causally-anchored multi-omic Mendelian Randomisation (MR) with graph-based deep learning for gene prioritisation in human exercise-ageing biology, using accelerometer-derived vigorous physical activity (VPA) in the UK Biobank as exposure. Combining multi-omic MR across five molecular layers, it asks whether causal inference and deep learning can recover exercise-responsive and potentially ageing-causal genes that were independently identified in prior studies. Enrichment for experimentally exercise-responsive genes was undetectable in the raw MR signal (p = 0.97) yet was recovered by the graph model (p = 0.007, reproducible across all initialisations); and the convergence between VPA MR-anchored and ageing-causal genes (significant on its own at 1.6-fold; p = 0.023) was likewise recovered by the graph model where p-value and effect-size ranking could not. The model further reproduced established acute exercise-responsive immune and lipid-metabolic programmes, supporting its recovery of genuine signal. Extending the prioritised genes to formal causal testing, systematic cis-MR with colocalisation across the eight convergent genes and four ageing outcomes identified cathepsin F (*CTSF*) as causally associated with exceptional longevity, with concordant positive estimates in the protein and LD-clumped expression arms and colocalisation support at the protein level. The contribution is therefore twofold: a model-free, like-for-like convergence between exercise-anchored and ageing-causal genes; and a graph-based method that recovers this convergence, together with exercise-responsive biology, beyond the reach of per-gene MR ranking.

Graphical abstract
This study introduces a causally-anchored deep learning approach to integrating multi-omic human data. The analytical pipeline begins with an exome-wide association study (ExWAS) and Mendelian randomisation (MR) with vigorous activity as exposure and omics markers as outcomes. MR serves as a causal anchor to prioritise genes from features derived across five molecular layers: CpG methylation, bulk and single-cell gene expression, protein levels, and glycan traits. These causally-anchored priors are unified within a supervised graph attention network (GAT) that integrates a protein-protein interaction network to learn a systems-level representation of the exercise-ageing axis. The model’s prioritised genes are then tested for statistical overlap with two independently derived reference sets: exercise-responsive genes (from an independent multi-omic human exercise study) and potentially ageing-causal genes (CpGs causal for ageing by epigenome-wide MR, mapped to genes). The analysis demonstrates that the graph model recovers enrichment for both reference sets that is not detectable in the raw MR signal. Created with Biorender.co

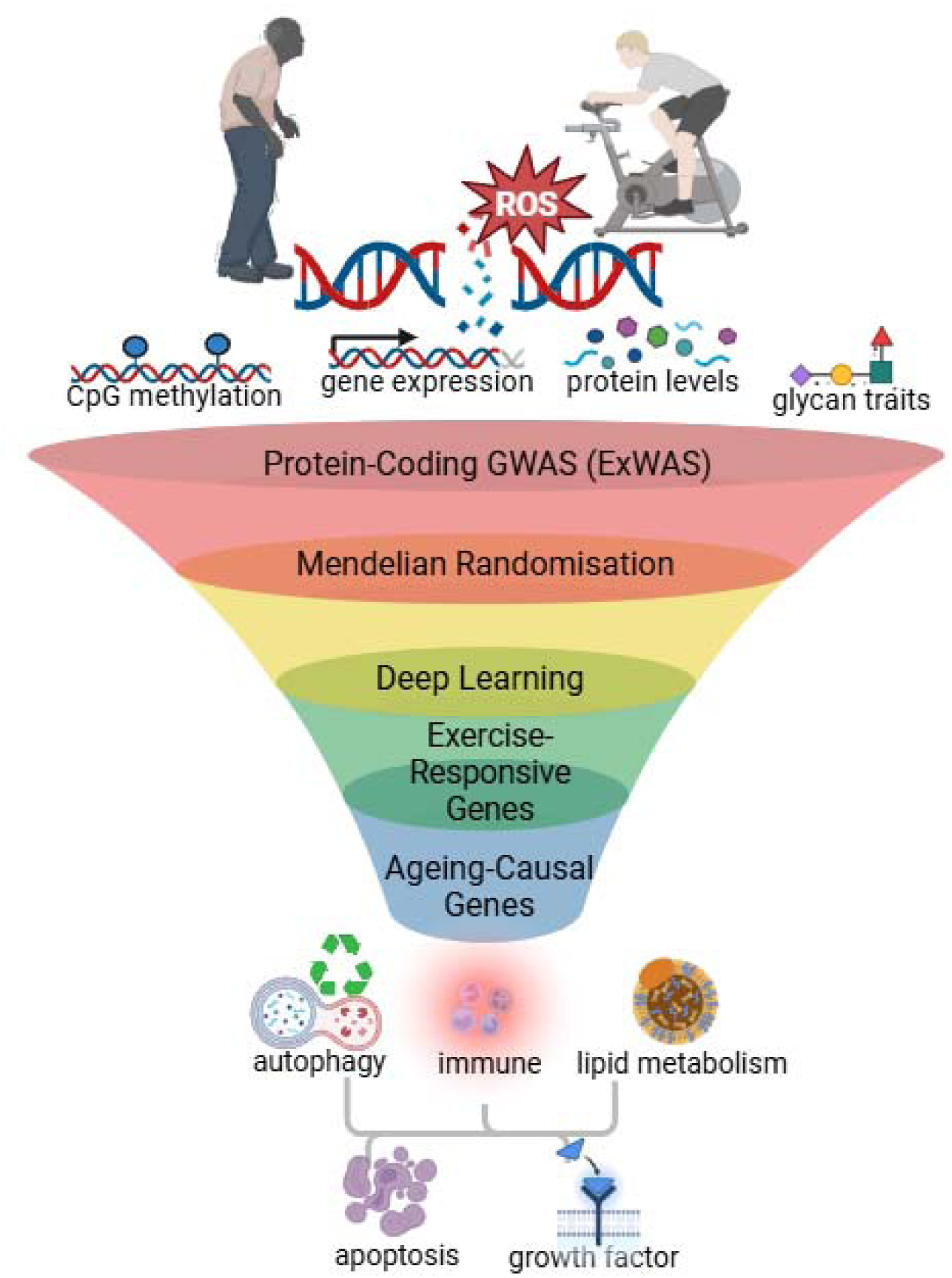

## Introduction

Regular physical activity is an effective behavioural intervention for preserving functional capacity and delaying the onset of age-related multimorbidity and mortality (Biswas et al., 2025). Yet, despite robust epidemiological evidence, the molecular pathways through which habitual exercise reconfigures proteomic, transcriptomic, epigenomic, and glycomic networks in humans remain incompletely understood. Dissecting these pathways is essential for moving beyond observational associations to mechanistic insight, and for identifying molecular regulators that could be targeted to mimic or potentiate the benefits of physical activity.

Large-scale genome-wide association studies (GWAS) in the UK Biobank (UKBB) have begun to map the genetic architecture of physical activity. Early efforts using self-reported and accelerometer-derived measures identified loci associated with habitual activity and sedentary behaviour (Klimentidis et al., 2018). Subsequent analyses of seven-day wrist accelerometry extended these findings to objectively measured activity, sleep, and circadian traits, identifying loci associated with overall movement and rhythmicity (Doherty et al., 2018). More recent multi-trait accelerometry GWAS further refined genetic signals across activity intensity, timing, and bout structure, using supervised algorithms to extract higher-resolution behavioural phenotypes from raw device data (Qi et al., 2022).

Importantly, device-based epidemiology now suggests that activity intensity is a key determinant of health. Using wrist-worn accelerometry in UKBB, Biswas et al. (2025) showed that as little as one minute per day of vigorous physical activity (VPA) was associated with health benefits compared to 10 or over 100 minutes of light-intensity activity, with outcome-specific equivalence estimates of approximately ∼90 minutes for diabetes risk, ∼70-85 minutes for cardiovascular disease risk, and >150 minutes for cancer outcomes. These findings reinforce a growing consensus that VPA carries disproportionate biological benefits, placing an emphasis and mechanistic focus on pathways that respond specifically to higher-intensity exercise exposures.

Despite increasingly refined behavioural phenotypes, translating polygenic physical activity signals into coherent molecular mechanisms remains challenging. Exercise-responsive pathways are dispersed across molecular layers and rarely converge through standard association-based prioritisation (Huang et al., 2025; MoTrPAC Study Group, 2024; Spence et al., 2025). This reflects broader limitations of GWAS translation, where loci are typically ranked by statistical significance rather than by biological efficiency or functional relevance. A recent appraisal of GWAS highlighted several structural barriers to translation, including reliance on additive heritability, proliferation of small effect associations, under-utilisation of multi-omic data, and the absence of frameworks that prioritise genes by how effectively they realise a trait (Huang et al., 2025). In response, Huang and colleagues introduced the concept of trait efficiency loci, arguing that the most informative regulators of complex traits may be those that mediate efficient structural and functional effects rather than those with the largest marginal effects. In parallel, Spence et al. (2025) demonstrated that conventional GWAS ranking strategies are systematically biased by gene length, pleiotropy, and stochastic variation, and formalised trait importance as a quantitative property of gene-trait relationships. Together, these advances motivate approaches that move beyond association to causally-anchored, structure and function-aware prioritisation of exercise-responsive molecular programmes (Huang et al., 2025; MoTrPAC Study Group, 2024; Spence et al., 2025).

This study builds on these conceptual advances by combining causal inference from Mendelian randomisation (MR) with a supervised graph attention network (GAT) to prioritise exercise-anchored genes across molecular layers. The model integrates gene-level causal MR evidence with the topology of a protein-protein interaction network to learn attention-weighted gene representations, organising sparse causal signal within its biological context rather than performing causal discovery or mechanistic modelling. By propagating causal evidence across the interaction network, this approach aims to resolve diffuse polygenic signal into a robust, reproducible set of exercise-ageing genes that are not recoverable from per-gene MR ranking alone. To situate these genes within ageing biology, the GAT-prioritised genes are tested for convergence with two independently derived reference sets: genes identified as causally implicated in biological ageing by epigenome-wide Mendelian randomisation (Ying et al., 2024), and genes identified as exercise-responsive in an independent multi-omic human exercise cohort (MoTrPAC Study Group, 2026). Crucially, the ageing reference is itself derived by causal MR rather than from correlative ageing clocks, providing a like-for-like causal comparison. Together, this framework of combining causal inference, supervised graph learning, and independent human experimentation provides a causally-anchored account of the genes through which VPA intersects with ageing biology, and illustrates how multi-omic graph learning can help bridge the long-standing gap between GWAS discovery and biological interpretation, while recovering signal not apparent at the level of individual associations.

## Results

### GAT recovers exercise-responsive and ageing-causal genes

To test whether the prioritised genes captured biology relevant to exercise and ageing, gene sets from each prioritisation method were assessed for enrichment of two independent, externally defined reference sets: genes identified as potentially causal for biological ageing derived by epigenome-wide MR (Ying et al., 2024), and genes experimentally responsive to acute exercise in an independent multi-omic human experiment (MoTrPAC Study Group, 2026). Enrichment was evaluated by hypergeometric overrepresentation: GAT rank-based methods against the connected protein-protein interaction STRING graph (n = 2,473 genes), and the model-free convergence between the two causal sets against the multi-omic MR universe (n = 2,959 MTI-scored genes).

At the gene-set level, the genes causally anchored to VPA (FDR < 0.05, n = 906) were significantly enriched for ageing-causal genes (16 observed versus 10.1 expected; 1.6-fold; p = 0.023), indicating a convergence between two independent MR analyses (one anchored on physical activity, the other on ageing traits) that was present prior to any deep learning step. This convergence was not, however, recovered by ranking genes on their MR statistics alone: neither best MR p-value nor multi-omic trait importance (MTI, effect-size based) produced significant ageing-causal gene enrichment at any ranking depth (p ≥ 0.31).

The supervised GAT, by contrast, concentrated the ageing-causal gene signal among its top-ranked genes and recovered significant enrichment robustly across the top 100 to 200 genes (p ≈ 0.02-0.024), an effect reproduced in five of five random initialisations. A supervised non-graph baseline (multilayer perceptron, MLP) recovered the enrichment only at the top 100 genes and lost significance as the ranking widened (p = 0.083 at K = 150; p = 0.180 at K = 200), localising the additional, ranking depth-robust signal specifically to the graph architecture.

The MoTrPAC exercise-responsive set behaved differently: No gene set or ranking method based on the MR signal alone showed any enrichment for experimentally exercise-responsive genes. The FDR-significant MR set (p = 0.97), p-value ranking (p = 0.97), effect-size ranking (p = 0.84), and the non-graph MLP (p = 0.12) were all non-significant, with the raw MR set marginally depleted. Only the GAT recovered a significant enrichment (44 observed versus 32 expected at the top 100 genes; p = 0.007), again reproduced in five of five initialisations, though this signal weakened as the ranking widened (p = 0.071 at K = 200). The undetectable enrichment at the level of individual MR associations emerged specifically through the graph model, indicating that network propagation surfaced exercise-responsive biology not represented in the per-gene multi-omic signal.

Taken together, these analyses establish two complementary results. First, a model-independent convergence between exercise-anchored and ageing-causal genes (p = 0.023) demonstrates that the multi-omic MR captures genuinely ageing-relevant biology. Second, the graph attention model recovered both this ageing-causal convergence and an exercise-responsive signal (MoTrPAC genes) that no ranking of the MR statistics (nor a non-graph supervised baseline, MLP) could recover, indicating that the contribution of the deep learning step lies not in re-ranking an already visible signal but in propagating sparse, distributed multi-omic evidence across the protein-protein interaction network to surface exercise-responsive and ageing-causal genes that per-gene analysis leaves latent. Table 1 summarises the results.

**Table 1.** Recovery of ageing-causal (Ying et al.) and acute exercise-responsive (MoTrPAC) genes by five prioritisation approaches. GAT rank-based methods were tested by hypergeometric overrepresentation against the connected STRING graph (n = 2,473 genes); the model-free MR-FDR convergence (top-left cell) was evaluated against the multi-omic MR universe (n = 2,959 MTI-scored genes). Ageing-causal gene set (Ying et al., 2024); exercise-responsive gene set (MoTrPAC Study Group, 2026). MLP multilayer perceptron; MTI multi-omic trait importance; GAT graph attention network.

| Method | Ying et al.<br>(chronic, $n=33$) | | | MoTrPAC<br>(acute, $n=948$) | | |
| --- | --- | --- | --- | --- | --- | --- |
|  | K=100 | K=150 | K=200 | K=100 | K=150 | K=200 |
| MR FDR set | **0.023** | — | — | 0.969 | — | — |
| MR p-value rank | 0.307 | 0.505 | 0.665 | 0.971 | 0.994 | 0.985 |
| MTI ($\beta$) rank | 0.307 | 0.505 | 0.665 | 0.839 | 0.793 | 0.953 |
| MLP (no graph) | **0.024** | 0.083 | 0.180 | 0.118 | 0.212 | 0.243 |
| GAT (graph) | **0.024** | **0.024** | **0.021** | **0.007** | **0.014** | **0.071** |

In the table above, values are enrichment p-values; K denotes the ranking depth, i.e. the top K highest-ranked genes from each method tested for overrepresentation of the reference set (top 100, 150, or 200); bold indicates p < 0.05. The MR, FDR-corrected row reports the single FDR-significant gene set (a fixed set, not a ranked list), so it has no K-dependent columns; ageing-causal (Ying et al.) gene enrichment is detectable in this raw set (1.6-fold, p = 0.023) and robustly recovered by the GAT across all ranking depths (5/5 initialisations), whereas the exercise-responsive (MoTrPAC) gene enrichment is undetectable in the raw MR set and all non-graph methods and is recovered only by the GAT (p = 0.007 at K = 100; 5/5 initialisations). Because MoTrPAC reflects the acute single-bout response, its enrichment supports broad exercise-responsiveness rather than chronic adaptation.

### Convergence across causal, experimental, and ageing-related evidence

To examine whether the genes implicated by multi-omic MR were supported by independent evidence, three gene sets were compared within the multi-omic MR universe (n = 2,959 MTI-scored genes): genes causally anchored to VPA (MR-anchored, FDR < 0.05; n = 906), genes experimentally responsive to acute vigorous exercise in the MoTrPAC cohort (n = 948), and potentially ageing-causal genes from Ying et al. (n = 33 in this universe). The two exercise/physical activity sets overlapped substantially, sharing 269 genes, consistent with partial concordance between genetically-anchored and experimentally-measured exercise biology. The ageing-causal gene set intersected both exercise/physical activity sets: 16 ageing-causal genes were VPA MR-anchored (a 1.6-fold enrichment over the 10.1 expected by chance at p = 0.023, hypergeometric), while 12 were MoTrPAC-responsive. Figure 1 summarises the overlap.

**Figure 1.**
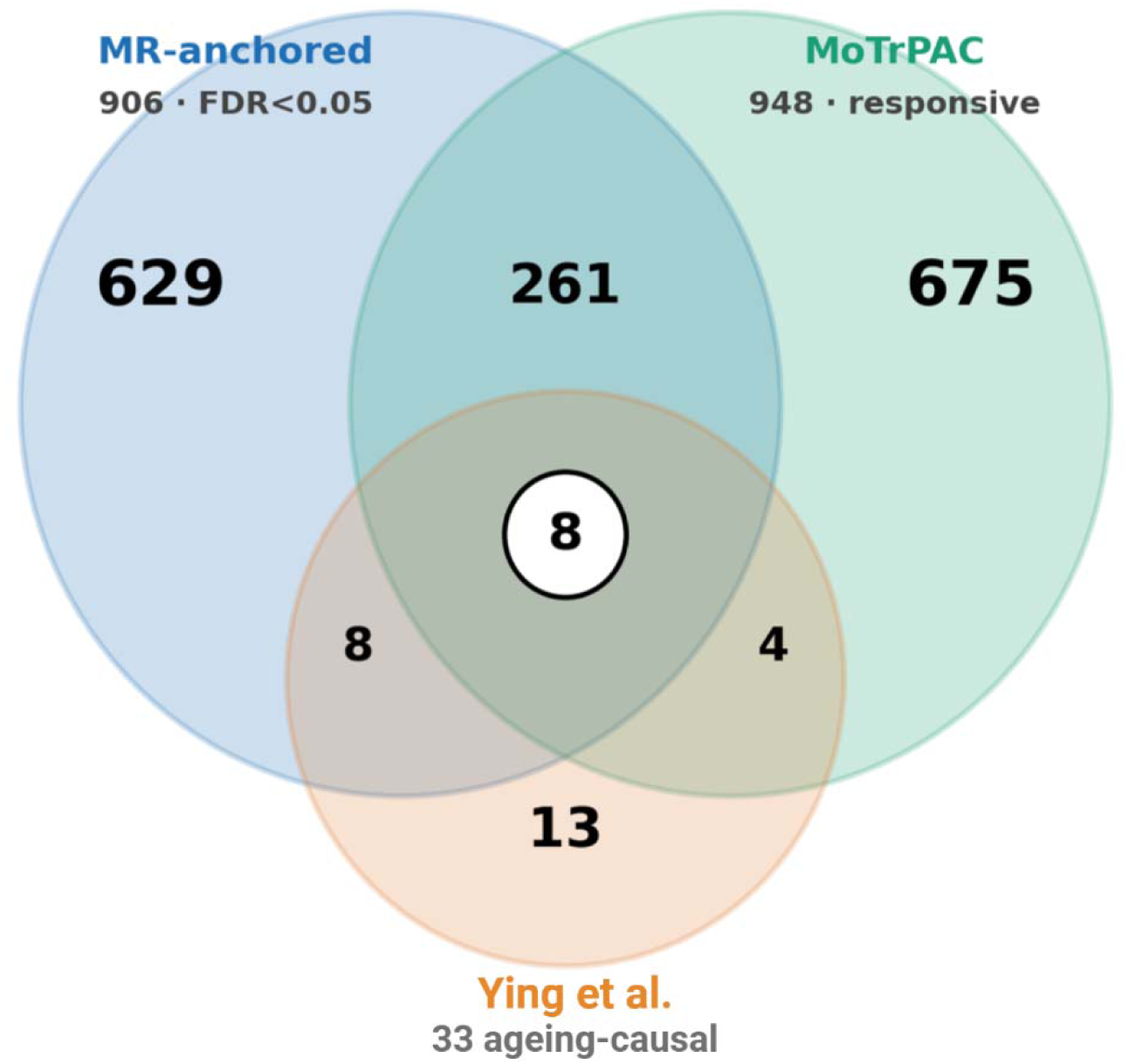
Overlap of three gene sets within the 2,959-gene multi-omic MR universe. Genes causally anchored to VPA by germline multi-omic MR (chronic exposure; n=906); genes responsive to a single acute exercise bout in MoTrPAC blood transcriptome and Olink proteome (acute response; n = 948); and genes potentially causal for biological ageing by germline MR (Ying et al.; chronic; n=33). Counts are per region; circle areas are not to scale. The MR-anchored and Ying et al. sets reflect chronic, germline-anchored exposures, whereas the MoTrPAC set reflects the acute transcriptional response to a single exercise bout; the MoTrPAC overlaps are therefore interpreted as exercise-responsiveness of the prioritised genes, not as validation of chronic adaptation.

**Figure 1.**
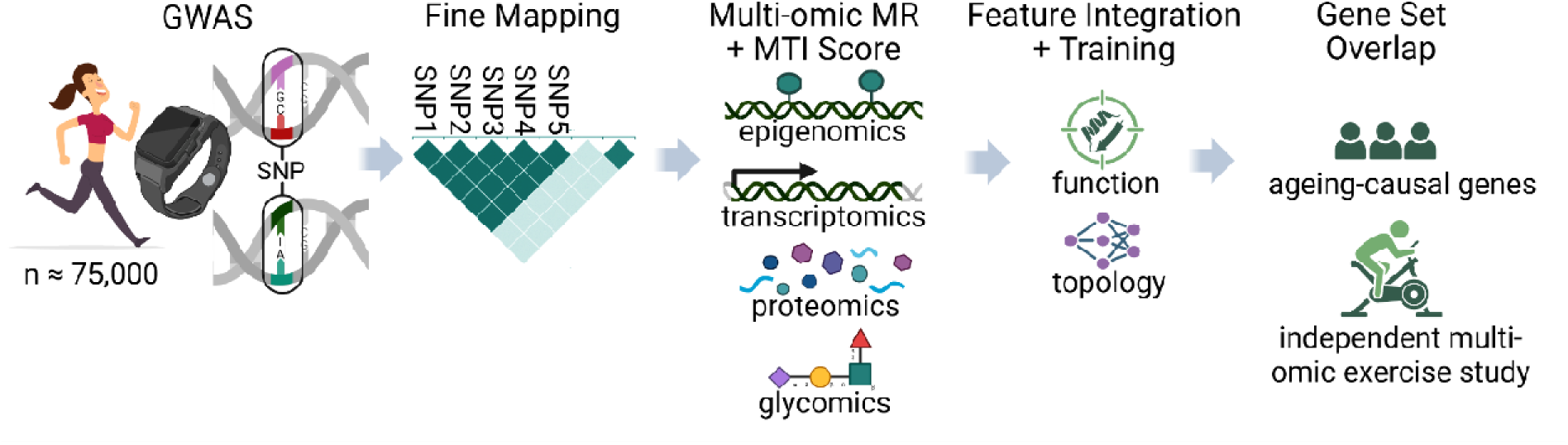
Summary of the analytical pipeline. GWAS, genome-wide association study; MR, Mendelian randomisation; MTI, multi-omic trait importance; SNP, single-nucleotide polymorphism. Created with Biorender.com.

Eight genes lay at the intersection of all three sets: *CTSD, CTSF, FADS1, FADS2, HEXIM1, IGFBP7, LTBP3, and RHOC*. These genes are simultaneously causally anchored to VPA, experimentally responsive to acute exercise, and potentially causal for biological ageing. Their biological annotations span lipid metabolism (*FADS1, FADS2*), autophagy and lysosomal proteostasis (*CTSD, CTSF*), growth factor and matrix signalling (*IGFBP7, LTBP3*), transcriptional and cardiac adaptation regulation (*HEXIM1*), and Rho-family cytoskeletal signalling (*RHOC*), programmes consistent with the systemic adaptations associated with habitual VPA. Two of the eight, *FADS2* and *HEXIM1*, were additionally ranked among the top 20 genes by the supervised GAT (ranks 3 and 18), linking the convergent evidence analysis to the model-based prioritisation.

Among acute exercise-responsive genes (MoTrPAC), those prioritised by the graph model (but not by MR alone) were strongly enriched for leukocyte activation and immune effector programmes. The graph model recovered significant enrichment of the MoTrPAC exercise-responsive set among its top-ranked genes (44 observed versus 32 expected at the top 100 genes; p = 0.007, reproducible across all five initialisations), a signal absent from the raw MR gene set (p = 0.97); and within these prioritised genes the immune/leukocyte activation programme was the dominant signal (169 enriched immune-related terms; best p ≈ 2 × 10⁻¹□; robust across all five initialisations). As this derives from the acute bout contrast, it most plausibly reflects transient immune mobilisation rather than chronic adaptation. The immune/leukocyte activation programme recovered by the graph model among MoTrPAC-responsive genes closely matches the dominant signal reported in MoTrPAC’s own whole blood analysis, in which the immediate response to acute exercise was dominated by immune pathways including NK and CD8⁺ T-cell, neutrophil, and Toll-like-receptor/cytokine signalling (MoTrPAC Study Group, 2026). This independent concordance supports the biological validity of the recovered programme. As the original authors note, however, whole-blood transcriptional changes cannot be fully attributed to within-cell regulation versus shifts in circulating immune cell proportions; the programme is therefore interpreted as the acute exercise-immune response (encompassing leukocyte mobilisation) rather than a chronic adaptive signature.

### Causal validation of prioritised genes against ageing outcomes

We tested the eight prioritised genes for causal association with four ageing outcomes (parental lifespan, healthspan, aging-GIP1, and exceptional longevity) using two-sample cis Mendelian randomisation with colocalisation, across two independent instrument layers: cis-pQTL (UKBB) for the six genes assayed on the Olink panel (*CTSD, CTSF, HEXIM1, IGFBP7, LTBP3, RHOC*), and cis-eQTL (eQTLGen) for all eight genes. Causal effects were estimated by inverse variance weighted (IVW) regression where two or more independent cis instruments were available, and by the Wald ratio otherwise; protein arm estimates used the lead cis-pQTL variant throughout; a gene-outcome pair was considered validated only if it showed FDR-significant MR, a Steiger-consistent direction, and colocalisation (conditional PP.H4 > 0.7). The MR Steiger test was applied to every gene-outcome signal, not only to validated pairs; for the non-validating genes the decisive failure was colocalisation rather than Steiger direction, as detailed below.

A single gene met all validation criteria: cathepsin F (*CTSF*) was causally associated with exceptional longevity. In the protein arm, genetically predicted higher *CTSF* was associated with increased odds of survival beyond the 90th percentile by the lead cis-pQTL variant (Wald β = 0.33; P = 1.8×10⁻³), with strong colocalisation (conditional PP.H4 = 0.78) and a Steiger-consistent direction. The expression arm reproduced this association in the same direction with multiple LD-clumped instruments (β = 0.19; P = 6.6×10⁻ □; six instruments; Cochran’s Q P = 0.99, no heterogeneity), with directionally concordant but sub-threshold colocalisation (conditional PP.H4 = 0.62). Colocalisation (the criterion discriminating a shared causal variant from confounding) meets the validation threshold at the protein level, with the expression arm providing heterogeneity-free supporting evidence. MR Steiger directionality for all FDR-significant gene-outcome pairs is reported in Supplementary Table 1; every pair was Steiger-consistent, so colocalisation was the criterion that distinguished validated from non-validated pairs.

*CTSF* showed a weaker association with aging-GIP1 (expression arm IVW β = 0.034, P = 5.0×10⁻□; protein-arm Wald P = 1.3×10⁻²), and null or inconsistent associations with healthspan and parental lifespan; none colocalised (conditional PP.H4 ≤ 0.30). Among the other genes, *FADS1* and *LTBP3* reached FDR-significant expression arm MR against aging-GIP1 without colocalisation (P = 1.0×10⁻³, FDR = 0.008, PP.H4 = 0.10; and P = 2.2×10⁻³, FDR = 0.013, PP.H4 = 0.33), while *FADS2* was nominally but not FDR-significant (P = 2.0×10⁻², FDR = 0.079). *HEXIM1* showed an apparent single-instrument association with aging-GIP1 (P = 7.9×10⁻□) that failed to colocalise in either arm (PP.H4 = 0.28/0.39). These illustrate the necessity of requiring colocalisation alongside MR.

Overall, of eight prioritised genes tested against four ageing outcomes across two instrument layers, one (CTSF) showed convergent, cross-layer causally-anchored and colocalisation evidence for exceptional longevity, while the majority of gene–outcome associations were null and several FDR-significant MR signals were resolved as non-colocalising. This pattern is consistent with causally-anchored prioritisation surfacing genuine, individually testable longevity biology, while demonstrating that set-level enrichment does not imply that every prioritised gene exerts an individually detectable causal effect on distal ageing phenotypes.

## Discussion

The central aim of this study was to derive a causally-anchored account of how VPA intersects with biological ageing, moving beyond per-gene associations to recover the genes through which the two converge. Integrating LD-aware, pleiotropy-filtered multi-omic MR with GAT learning produced three principal findings. First, a convergence between genes causally anchored to VPA and genes causally implicated in ageing was detectable directly in the MR output, before any deep learning step. The set of genes causally anchored to VPA (FDR < 0.05) was enriched 1.6-fold for the ageing-causal reference genes of Ying et al. (16 observed versus 10.1 expected; p = 0.023). Because both sets are derived by germline-instrumented, pleiotropy-filtered MR, this is a like-for-like convergence of two methodologically matched analyses, present in the raw MR signal and requiring no machine learning to detect.

Second, the supervised GAT recovered both this ageing-causal enrichment and an exercise-responsive (MoTrPAC) enrichment that no ranking of the MR statistics, and no non-graph supervised baseline (MLP), could recover, localising the deep learning contribution to the propagation of distributed causal evidence across the protein-protein interaction network rather than to the re-ranking of an already visible signal. Third, eight genes emerged at the intersection of all three lines of evidence, being simultaneously causally anchored to VPA, responsive to acute exercise, and potentially causal for biological ageing. These genes concentrate metabolic, proteostatic, immune, and growth-factor signalling functions and recur across independent multi-omic studies of exercise, as discussed below.

The convergence between exercise-anchored and ageing-causal genes is the most epistemically robust finding of this study, because of how the two sets were derived. Conventional attempts to connect exercise to “biological age” rely on epigenetic clocks trained to maximise correlation with chronological age or mortality; as Ying et al. (2024) showed by epigenome-wide MR, none of the major clocks (Horvath, Hannum, PhenoAge, GrimAge) are enriched for CpG sites with causal effects on ageing-related traits. A convergence between exercise genes and a clock signature would therefore be a convergence with a correlative biomarker of uncertain causal meaning. The convergence reported here is of a stronger kind: both sides are derived under the same causal framework (germline instruments, two-sample MR, and MR-Egger pleiotropy filtering) with the ageing side instrumenting DNA methylation against eight lifespan and healthspan traits using the same GoDMC methylation quantitative trait loci and the same Egger-based control. The shared genes are thus not merely co-correlated with age and activity but independently implicated, by germline instrumentation, as potentially causal for both, arguing that habitual VPA engages genes lying on the causal architecture of human ageing rather than only genes that respond to it.

The magnitude should nonetheless not be overstated. The ageing-causal reference comprises only 33 testable genes within the analysed universe, so the 16-gene overlap, while significantly above the chance expectation of approximately ten, rests on modest numbers. The convergence was therefore characterised by per-gene functional annotation rather than pathway-enrichment testing, which would be underpowered at this size; the biological coherence of the overlapping genes provides qualitative support, but the central claim is statistical: two independently constructed, potentially causal gene sets share more genes than expected by chance, before any modelling step intervenes.

The second finding concerns what the graph model adds beyond the raw causal signal. A persistent difficulty in translating polygenic signals into mechanism is that the relevant genes are dispersed across molecular layers and rarely converge through association-based ranking, which orders loci by statistical significance rather than by position within a functional network. The results make this concrete. The MoTrPAC exercise-responsive set showed no enrichment in the MR-only output: neither the FDR-significant set (p = 0.97), nor p-value ranking (p = 0.97), nor effect-size ranking (p = 0.84) and was, if anything, marginally depleted; a non-graph MLP trained on the same node features also failed (p = 0.12). Only the GAT recovered the signal robustly: 44 observed versus 32 expected among its top 100 genes (p = 0.007), reproduced in five of five random initialisations. For the ageing-causal set the pattern was parallel: the MLP recovered enrichment only at the top 100 genes and lost it as the ranking widened (p = 0.083 at K = 150; p = 0.180 at K = 200), whereas the GAT held significance across the top 100 to 200 genes in every initialisation.

The interpretation of this contribution must be carefully bounded. Because the model’s supervision target (the MTI score) is itself derived from the MR effect sizes, the network performs no independent causal discovery: it cannot generate causal information that the upstream MR did not contain, and its attention weights are indicators of relative importance within the learned representation, not estimates of biological mechanism. What it does is propagate the sparse, distributed causal signal across the protein-protein interaction graph, so that genes embedded among many causally-implicated neighbours are surfaced even when their own per-gene signal is weak. That an enrichment undetectable at the level of individual associations emerges specifically through graph propagation (and not through an MLP with identical inputs) indicates that exercise-responsive biology is encoded in the topology of the causal signal, in which genes connect to which, rather than in the marginal strength of any single gene. This is consistent with the polygenic, network-distributed character of complex-trait biology articulated in recent appraisals of GWAS translation (Huang et al., 2025; Spence et al., 2025), and with the demonstration that message-passing architectures recover predictive structure from topology combined with continuous biological attributes (Yang et al., 2025; Vinyard et al., 2025). The contribution of the deep learning step is thus denoising and propagation, not discovery, a distinction the architecture was explicitly designed to respect.

Two analyses underpin the interpretation that follows, and must be kept separate. Pathway enrichment testing was applied to the broad set of GAT-prioritised genes (the top-ranked ∼100-200), large enough to support hypergeometric testing; this is the source of all programme-level statistics. Functional annotation was applied to the eight triple-convergent genes, which are too few for enrichment and are interpreted gene by gene. The programme-level findings below therefore derive from the prioritised gene set, not from the eight convergent genes.

Among the prioritised genes, the signal the graph model surfaced most strongly was a leukocyte activation and immune-effector programme, robust across all five initialisations and independently corroborated: the dominant transcriptional response in MoTrPAC’s own whole-blood analysis was an immune signature spanning natural-killer and CD8⁺ T-cell, neutrophil, and Toll-like-receptor and cytokine signalling (MoTrPAC Study Group, 2026). That a model trained only on causal multi-omic effect sizes, with no immune annotation supplied, recovers the same programme the experimental data independently report lends external support to the biological validity of the recovered signal. Two caveats apply: The MoTrPAC contrast reflects the immediate response to a single moderate-to-vigorous endurance bout, not chronic adaptation, so the enrichment indexes exercise-responsiveness in general rather than the durable remodelling repeated training produces; and, as the original authors emphasise, whole blood transcriptional changes after acute exercise cannot be fully separated from shifts in the proportions of circulating immune cells, since vigorous exercise acutely mobilises leukocytes. The programme is accordingly interpreted as the acute exercise-immune response, including leukocyte mobilisation, rather than a chronic adaptive signature.

The eight triple-convergent genes (*CTSD, CTSF, FADS1, FADS2, HEXIM1, IGFBP7, LTBP3, RHOC*) are too few to support enrichment testing and are interpreted individually, their established biology examined for coherent mechanistic threads. These threads are advanced as hypotheses consistent with known gene function, not as relationships demonstrated in this work. The first thread links lipid biosynthesis to inflammation resolution, and has independent statistical support: a lipid-metabolic signal was robustly enriched among the prioritised genes across all five initialisations (long-chain fatty-acid binding, FDR = 0.003; fatty-acid binding, FDR = 0.005; response to lipid, FDR = 0.011). *FADS1* and *FADS2* encode the Δ5 and Δ6 desaturases that are rate-limiting in polyunsaturated fatty-acid biosynthesis, generating the long-chain precursors of both pro-inflammatory eicosanoids and specialised pro-resolving mediators such as resolvins and protectins (Serhan, 2014); variation at the *FADS* locus is among the strongest known genetic determinants of circulating polyunsaturated fatty-acid and lipid traits (Schaeffer et al., 2006). Because resolution of an inflammatory episode depends on the timely production of these mediators, FADS-controlled desaturase activity provides a candidate bridge between lipid metabolism and the acute exercise-immune programme (the same desaturases that shape membrane and signalling-lipid composition also gate the lipid mediators that terminate an inflammatory response) and thus a candidate coupling point between the two robust programmes the model recovered rather than a merely parallel signal.

The second thread links lysosomal proteostasis to lipid handling. *CTSD* and *CTSF* encode lysosomal proteases (an aspartyl and a cysteine cathepsin, respectively) central to autophagic-lysosomal degradation; loss-of-function in either causes neuronal ceroid lipofuscinosis, a lysosomal storage disorder marked by accumulation of lipofuscin, itself a recognised hallmark of ageing (Siintola et al., 2006; Smith et al., 2013; López-Otín et al., 2023). Lysosomal function and lipid metabolism intersect through lipophagy and the lysosomal turnover of lipid-laden material (Singh et al., 2009), such that declining cathepsin-dependent proteostatic capacity and altered lipid composition could act on a shared membrane and organelle substrate. Two lysosomal cathepsins among the convergent genes is, at the level of individual gene biology, consistent with the centrality of proteostatic maintenance to ageing.

The third thread links growth factor and matrix signalling to cellular senescence. *IGFBP7* is an established inducer and marker of cellular senescence, acting through autocrine modulation of insulin-like growth factor signalling (Wajapeyee et al., 2008), and is a component of the senescence-associated secretory phenotype. *LTBP3* regulates the bioavailability and activation of transforming growth factor-β by sequestering its latent complex within the extracellular matrix (Robertson et al., 2015); TGF-β in turn governs matrix remodelling, fibrosis, and the senescence-associated secretory programme, and modulates immune tone. Together they implicate a senescence-matrix axis (both cellular senescence and altered intercellular communication being canonical hallmarks of ageing; López-Otín et al., 2023) through which habitual VPA might influence the tissue micro-environment, with IGF-TGF-β cross-talk a plausible coordination point.

The fourth thread connects the set to the immune programme through cytoskeletal and transcriptional regulation. *RHOC* encodes a Rho-family GTPase that drives actomyosin remodelling and directed cell migration; in immune cells, Rho-GTPase-dependent cytoskeletal reorganisation is required for leukocyte motility and trans-endothelial migration (Heasman & Ridley, 2008), making *RHOC* a mechanistic link to the leukocyte mobilisation component of the recovered immune programme. *HEXIM1*, as the principal inhibitor of the positive transcription elongation factor P-TEFb, regulates RNA-polymerase-II pause-release and thereby the breadth of inducible transcriptional responses, including stress and inflammation-responsive programmes (Michels & Bensaude, 2018), a coherent member of a gene set responding to a systemic physiological stressor.

The fifth thread connects the set to the oxidative damage and repair axis. Vigorous exercise transiently elevates reactive oxygen species (ROS), which act not only as potential damage agents but as adaptive signalling inputs to redox-sensitive stress response programmes (the mitohormetic view of exercise; Ristow & Schmeisser, 2011; Williamson et al., 2020). The lysosomal cathepsins *CTSD* and *CTSF* clear oxidatively modified and cross-linked macromolecules (lipofuscin, the storage material accumulating when these proteases fail, being itself an oxidised aggregate; Höhn & Grune, 2013), and the desaturases *FADS1* and *FADS2* supply the polyunsaturated fatty acids that are the principal substrate for ROS-driven lipid peroxidation, linking redox state to membrane and signalling-lipid composition.

Taken together, the eight genes are not a disparate list but plausibly interlocking components of a metabolic-proteostatic-immune signalling network: the desaturases *FADS1* and *FADS2* coupling metabolism to inflammation resolution; the lysosomal cathepsins *CTSD* and *CTSF* maintaining proteostasis on a shared lipid-membrane substrate; *IGFBP7* and *LTBP3* regulating senescence and matrix tone; *RHOC* enabling the cytoskeletal basis of immune mobilisation; *HEXIM1* gating the transcriptional response; and the cathepsins and desaturases together intersecting the oxidative damage axis. These connections are inferential, drawn from established gene biology rather than tested here; the threads are advanced as mechanistic hypotheses for future interrogation, particularly the FADS-resolution and RHOC-migration bridges to the immune programme.

The programmes recovered here are not idiosyncratic to this analysis but recur across the major independent multi-omic studies of exercise, lending external support to the biological coherence of the prioritised genes despite the methodological differences between approaches. The immune signal is the most strongly corroborated. In the Molecular Transducers of Physical Activity Consortium’s whole-organism map of endurance training across 19 rat tissues, the adaptive response was characterised by widespread regulation of immune, metabolic, stress-response and mitochondrial pathways (MoTrPAC Study Group, 2024), while the same consortium’s human acute study (the exercise-responsive reference used here) reported an immediate response dominated by immune programmes. That a leukocyte activation programme is the dominant exercise signature in both a multi-tissue animal training study and an acute human study, and that the graph model recovers precisely this programme from causal multi-omic evidence alone, indicates that the immune component of the exercise response is robust across species and analytical frameworks; the concordance with the chronic training data suggests immune regulation is a feature of both the acute and the trained states.

The lipid-metabolic signal is similarly concordant, and here the dedicated mitochondrial analysis of the MoTrPAC rat data is especially pertinent. Amar et al. (2024) integrated the transcriptome, proteome, and post-translational modifications across trained rat tissues to resolve the mitochondrial response to endurance training, finding mitochondrial adaptation to be a central, coordinated, and cross-tissue feature of exercise, apparent at the protein and post-translational level rather than the transcriptome alone, with lipid and fatty-acid metabolism tightly coupled to mitochondrial remodelling. This bears on the present findings in two respects. It reinforces the biological centrality of the lipid-metabolic programme recovered here, including *FADS1* and *FADS2*, since fatty acid handling and β-oxidation are core to the mitochondrial substrate metabolism Amar et al. identify as a dominant axis of the trained response. And it underscores a limitation of any blood and transcript-anchored analysis: because the mitochondrial signal Amar et al. resolved is carried by the proteome and post-translational modifications in metabolically active tissues such as muscle and heart, a peripheral-blood, MR-anchored framework such as the present one would be expected to capture the metabolic dimension only partially, through its lipid-handling component, rather than as a full mitochondrial programme. The robust recovery of a lipid-metabolic programme here is therefore best interpreted as a blood-accessible window onto the lipid-mitochondrial-metabolic axis these tissue-resolved studies place at the centre of the exercise response.

The growth factor and matrix component of the convergent set finds its closest independent parallel in recent multi-omic work on exercise and brain ageing. Li et al. (2025), profiling single-nucleus transcriptomes and chromatin accessibility in an Alzheimer’s mouse model after prolonged voluntary exercise, found that the protective transcriptional networks converged specifically on growth factor signalling (epidermal growth factor receptor and insulin signalling), with the cognitive benefits of exercise abolished by pharmacological inhibition of this pathway. Two of the eight convergent genes identified here, *IGFBP7* and *LTBP3*, are regulators of IGF and TGF-β signalling respectively, and the independent identification of growth factor signalling as the node on which exercise’s protective effects converge in a different species, tissue, and experimental paradigm strengthens the case that growth factor and matrix regulation is a genuine axis of the exercise-ageing relationship. That four independent multi-omic exercise studies (the multi-tissue animal training atlas of the MoTrPAC Study Group, the cross-tissue mitochondrial analysis of Amar et al., the human acute study of the MoTrPAC Study Group, and the brain-ageing intervention of Li et al.) converge on the same broad programmes the GAT recovers is meaningful external corroboration: the genes prioritised by causally-anchored graph learning occupy the same biological space that direct experimental profiling of the exercise response repeatedly identifies, while the present approach adds a specifically causal dimension, anchoring these programmes to germline-instrumented physical activity and to genes independently implicated in the causal architecture of ageing.

A further line of concordance comes not from molecular profiling but from the genetic architecture of physical activity itself. In the largest GWAS of physical activity to date, Wang et al. (2022) combined up to 703,901 individuals across 51 studies in a multi-ancestry meta-analysis, identifying 99 loci associated with moderate-to-vigorous physical activity, leisure screen time, and sedentary behaviour, with the implicated genes pointing predominantly to brain and, to a lesser extent, skeletal muscle biology. This reinforces the present work in two respects. It confirms that habitual physical activity is a heritable, polygenic trait with a tractable genetic architecture, the premise on which the germline-anchored MR instrument depends. And its MR analyses showed that the apparent beneficial effects of higher physical activity and lower sedentary behaviour on cardiometabolic risk factors and disease were largely mediated or confounded by body mass index, providing independent justification for treating BMI as a central covariate in the ExWAS and underlining that physical activity and body composition are causally entangled, so the activity-anchored signal recovered here is best interpreted as the component of vigorous activity at least partially separable from its effects on, and shared genetic basis with, body mass. That entanglement also means the BMI adjustment carries a recognised risk of attenuating genuine activity signal operating through body composition, a tension revisited among the limitations. Consistent with physical activity being a behaviour, Wang et al. located its genetic architecture predominantly in the brain, a compartment the present blood-anchored analysis does not sample, framing the circulating molecular programmes recovered here as downstream of, rather than coextensive with, the neural architecture that governs activity behaviour itself.

The single gene to show convergent, cross-layer causal evidence in this analysis was *CTSF*, biologically coherent as a longevity-associated target. *CTSF* encodes a lysosomal cysteine protease whose loss-of-function mutations cause type B Kufs disease, an adult-onset neuronal ceroid lipofuscinosis characterised by defective lysosomal degradation and accumulation of storage material (Smith et al., 2013); declining lysosomal and autophagic capacity is a recognised hallmark of ageing (López-Otín et al., 2023). The direction of the estimate (higher genetically predicted *CTSF* associated with greater odds of exceptional longevity, consistent across protein and expression instruments) is mechanistically plausible under a model in which preserved lysosomal proteolytic capacity supports the maintenance of proteostasis into later life, aligning with the broader finding that exercise-responsive, causally-anchored prioritisation recovered genes concentrated in proteostatic and metabolic maintenance rather than in any single canonical longevity pathway (two of the eight convergent genes, *CTSD* and *CTSF*, being lysosomal cathepsins).

Across both molecular layers the cis-MR estimates were positive and directionally consistent: in the LD-clumped expression arm, multi-instrument IVW gave β = 0.19 (P = 6.6 × 10⁻) with no between-instrument heterogeneity (Cochran’s Q P = 0.99), and in the protein arm the single independent cis instrument gave a concordant Wald estimate β = 0.33 (P = 1.8 × 10⁻³). Colocalisation was the discriminating criterion and was asymmetric between layers: the protein-level signal exceeded the threshold (conditional PP.H4 = 0.78) whereas the expression-level signal did not (0.62). The protein-level analysis therefore carries the primary colocalisation evidence on which *CTSF* meets the validation criteria, while the expression layer provides directionally concordant, heterogeneity-free support. That the headline cis-MR associations are nominal (P ≈ 10⁻³) rather than genome-wide reflects the conservative LD clumping applied; colocalisation, not MR p-value magnitude, distinguishes a shared causal variant from confounding.

Several limitations bound the CTSF interpretation. The estimate is a cis-regulatory MR association and should be read as evidence that genetically determined *CTSF* abundance is associated with longevity, not that pharmacological modulation of *CTSF* would extend lifespan; these are distinct claims, separated by the gap between a validated genetic association and a validated drug target. The longevity GWAS is modestly powered (11,262 cases), and although protein-level colocalisation exceeded threshold the expression-level did not, leaving residual uncertainty about a shared causal variant. Instruments were LD-clumped (r² < 0.001, 10 Mb) against the 1000 Genomes European panel, with the protein arm represented by the single lead cis-pQTL variant via the Wald ratio, yielding estimates more conservative than distance-based pruning would; and instruments derive from blood-based pQTL and eQTL resources, so tissue-specific effects in lysosomally active tissues may be incompletely captured. *CTSF* was a single colocalising signal among many gene-outcome pairs tested and warrants replication in independent longevity cohorts and, ideally, experimental follow-up before any causal claim is considered established. It is therefore presented as a prioritised, cross-layer-supported candidate for exceptional longevity rather than a confirmed determinant of human lifespan.

Several features of the analysis bear on how much confidence the recovered signals can carry. The final GAT rests on multi-omic causal evidence and network topology alone: structural and functional annotations were removed as node features, so the model could not recover immune or lipid enrichments simply by being told in advance which genes carry such annotations: the recovered enrichments reflect signal propagated through the multi-omic effect sizes and the interaction network, not a re-reading of pre-supplied annotation. The supervision target was protected against information leakage: because the MTI score is itself computed from the per-layer MR effect sizes that also serve as the model’s input features, an explicit leakage guard excluded every MTI-derived column from the inputs and asserted their absence before training, so the model had access to the individual per-layer effect sizes but never to the composite quantity it was asked to predict, and any predictive performance reflects genuine learning over the graph rather than self-copying. The recoveries were tested for stability rather than reported from a single run (across ranking depths top 100, 150, 200) and five independent initialisations, with significance required in the majority of seeds; the immune and lipid programmes met this in all five. Most importantly, the central causally-anchored claim of this study does not depend on the deep learning model at all: the convergence between exercise-anchored and ageing-causal genes was established model-free, directly between the two MR gene sets, before any graph-learning step. The model’s role is to demonstrate that this convergence (and the related exercise-responsiveness signal invisible to per-gene ranking) can be recovered and concentrated among prioritised genes, not to generate the underlying convergence itself.

Several limitations bound the conclusions. The most pervasive is ancestry: every exposure instrument and every outcome quantitative-trait-locus dataset used here derives from European ancestry samples, so the convergence and the prioritised genes apply most directly to European-ancestry populations, and extending the framework to multi-ancestry data is a priority both for equity and for the robustness of the causal inferences. A second concerns tissue: all outcome layers and the exercise-responsive reference are blood-accessible, so the convergence is most directly interpretable for blood-accessible molecular processes, and its extension to skeletal muscle, adipose, vascular, or neural tissue requires tissue-specific data not available here; plasma proteomics is moreover biased toward secreted and circulating factors and under-represents intracellular regulators. A third is intrinsic to MR: of its three core assumptions only relevance is fully verifiable, while independence and exclusion restriction can be supported and tested but not proven; the MR-Egger intercept tests directional but not balanced pleiotropy, and the LD-aware and overlap-aware corrections mitigate but do not eliminate bias from correlated instruments or sample overlap. The exposure phenotype, derived from accelerometer-measured vigorous activity in a sub-sample that consented to wear a monitor, is also subject to selection bias plausibly correlated with the behavioural intentionality the phenotype captures. For these reasons the estimates are described throughout as causally-anchored rather than definitively causal. A fourth concerns the reference sets: the ageing-causal set is small (33 testable genes), so the convergence, while statistically robust, rests on modest numbers and was characterised by per-gene functional annotation rather than pathway enrichment testing; the exercise-responsive reference reflects a single acute vigorous endurance bout whereas the UK Biobank exposure is a germline-anchored proxy for habitual, lifelong vigorous activity, so their overlap indicates broad exercise-responsiveness of the prioritised genes rather than validation of the chronic activity response. Finally, the convergence is a convergence, not a demonstration of mediation: it indicates that these gene sets overlap more than chance, not that physical activity acts on ageing through these specific genes, which would require explicit mediation analysis or interventional data.

Several lines of work follow directly from these findings and limitations. The most immediate is the move from causal anchoring toward interventional causal inference: the present framework identifies genes whose germline liability links exercise and ageing but cannot establish that perturbing them alters the exercise response. Datasets in which gene expression is experimentally perturbed (CRISPR or pharmacological screens with multi-omic readouts) would allow the convergent genes, and particularly the eight triple-convergent candidates, to be tested for causal mediation directly, transforming the model from a denoising tool into one capable of genuine causal discovery. A second direction is the experimental interrogation of the specific mechanistic hypotheses nominated here: the FADS-controlled lipid-mediator bridge between metabolism and inflammation resolution, and the RHOC-dependent cytoskeletal basis of leukocyte mobilisation, are concrete, testable links examinable through lipidomic profiling of pro-resolving mediators and immune-cell migration assays across an acute exercise time course; the growth-factor axis nominated by *IGFBP7* and *LTBP3*, and independently identified by Li et al. (2025), through targeted perturbation of IGF and TGF-β signalling; and the oxidative damage thread by pairing damage-repair assays with *CTSD, CTSF, FADS1* and *FADS2*. A third is the extension of the framework across ancestries and tissues, addressing the two most pervasive limitations simultaneously as multi-ancestry GWAS and tissue-resolved quantitative-trait-locus resources become available. The distinction between correlative and causal markers of ageing, central to the interpretation here, is itself an area of rapid methodological development: beyond the epigenome-wide causal framework of Ying et al. (2024), recent work resolves ageing signatures at spatial and single-cell resolution (e.g. spatial transcriptomic clocks that reveal cell-proximity effects in brain ageing; Sun et al., 2025), and integrating such spatially and causally-resolved ageing references with the exercise-anchored framework would allow the convergence to be examined not only at the level of which genes are shared but of where, and in which cellular neighbourhoods, that convergence is realised. Finally, the relationship between these molecular findings and emerging mechanistic-demographic models of ageing merits exploration: models conceptualising ageing as the stochastic accumulation of cellular damage governed by tightly constrained production and removal dynamics, with lifestyle exposures modulating damage tolerance and overall robustness (Shenhar et al., 2025, preprint), are agnostic to molecular identity, and the gene-level resolution provided here offers candidate substrates through which exercise might implement those robustness mechanisms, a formal integration of the molecular and demographic scales being a promising avenue for connecting the two accounts of how physical activity shapes ageing.

## Conclusion

This study set out to test whether the genes through which VPA acts converge with the genes causally implicated in ageing, and whether a graph-based model could recover that convergence from sparse, distributed multi-omic causal evidence. A model-independent convergence between exercise-anchored and ageing-causal genes (1.6-fold; p = 0.023), established by two methodologically matched MR analyses, demonstrates that habitual VPA engages genes lying on the causal architecture of human ageing, concentrated in metabolic, proteostatic, immune, and growth factor signalling biology. Building on this, a supervised GAT recovered both the ageing-causal convergence and an acute exercise-immune programme that neither per-gene MR nor a non-graph supervised baseline could detect, localising the model’s value to the propagation of distributed causal signal across the protein-protein interaction network rather than to the re-ranking of an already-visible one.

Eight genes (*CTSD, CTSF, FADS1, FADS2, HEXIM1, IGFBP7, LTBP3, RHOC*) emerged as simultaneously VPA-causal, exercise-responsive, and ageing-causal, their biology cohering into a candidate metabolic-proteostatic-immune signalling network whose specific mechanistic links are now nominated for experimental test. These programmes are precisely those that the major independent multi-omic studies of exercise repeatedly identify: from the multi-tissue animal training atlas of the Molecular Transducers of Physical Activity Consortium (2024) and its mitochondrial analysis (Amar et al., 2024), to the human acute exercise response (MoTrPAC Study Group, 2026), to the growth-factor network underlying exercise’s protective effects in the ageing brain (Li et al., 2025). That a causally-anchored, network-level prioritisation recovers the same biological space that direct experimental profiling identifies, while adding a specifically causal dimension absent from those descriptive maps, is the central contribution of this work.

This study also reframes the role of computational multi-omics in exercise-ageing biology. The model does not generate mechanism; its supervision target is itself causally derived, and its honest function is to recover, from diffuse genetic evidence, the convergent genes that warrant experimental interrogation, and to state clearly where that convergence does and does not extend. In moving from association to causally-anchored, network-level prioritisation, and in grounding its findings in convergence with both an independently causally-anchored ageing reference and the experimentally-derived programmes of the wider field, this work offers a reproducible and interpretable means of translating the diffuse polygenic signal of physical activity into a defined, testable set of exercise-ageing genes, with a transparent account of the boundary between what the approach can causally anchor and what remains for experiment to establish.

## Methods

### GNN Model Overview

The analysis proceeded in five stages: 1) an exome-wide association study (ExWAS) identified protein-coding variants associated with habitual vigorous physical activity (VPA), providing genetic instruments; 2) Linkage disequilibrium (LD)-aware, overlap-aware, pleiotropy-filtered Mendelian randomisation (MR) estimated the causal effect of VPA on each gene across five molecular layers; 3) these gene-level MR effects were aggregated into a continuous multi-omic trait importance (MTI) score; 4) a supervised graph-attention network (GAT) propagated the MTI signal over a protein-protein interaction network to produce a denoised gene ranking; and 5) the ranked genes were tested for statistical overlap with independently defined exercise-responsive and ageing-causal gene sets.

Graph neural networks are well suited to modelling biological systems in which functional effects emerge from structured interactions among entities rather than from isolated features. Recent advances in deep learning have demonstrated that appropriately constrained neural models can recover interpretable, biologically meaningful regulators from high-dimensional molecular data, including through generative dynamical frameworks that learn continuous biological processes from transcriptomic measurements (Vinyard et al., 2025). Complementary work in geometric deep learning has further shown that message-passing architectures integrating topology with continuous, biologically meaningful attributes can learn predictive representations of complex biological systems (Yang et al., 2025). Motivated by these advances, a GAT framework was used to propagate causally-anchored and topological information across a protein-protein interaction graph, capturing higher-order relationships between genes that are not evident from marginal MR or single-omic analyses alone.

The supervised GAT functions as a representation learning and denoising model: because its supervision target (the MTI score) is derived from MR outputs, the model does not constitute an independent causal discovery engine but learns graph-informed embeddings that stabilise and de-noise MR-derived gene scores. It is not interpreted as a mechanistic model of biological regulation, and attention weights are treated as context-dependent indicators of relative importance rather than as evidence of causal mechanism. Figure 1 summarises the model.

### Datasets

#### Exposure (physical activity)

Genetic instruments for habitual VPA were derived from UKBB, a prospective cohort of approximately 500,000 middle-aged and older participants with deep phenotypic and genotypic data. Exposure SNPs were generated from ExWAS (protein-coding GWAS) using the UKBB Oxford Quality-Controlled Functional Equivalence (OQFE) whole-exome sequencing (WES) data, restricted to the subset of approximately 75,000 participants with seven-day wrist-worn accelerometry data.

This accelerometry subset was retained in full, and participants were not excluded on the basis of prevalent disease. Restricting instrument discovery to ostensibly healthy individuals would condition the sample on health status, which is both a downstream consequence of habitual physical activity and a correlate of the ageing-related outcomes under study; conditioning on such a variable is conditioning on a collider and can induce selection bias, opening spurious pathways between the genetic instruments and the outcome that distort the genotype-exposure associations anchoring the downstream MR. This concern is not hypothetical for physical activity, where low activity is simultaneously a cause and a consequence of poor health (the “sick-quitter” or reverse-causation effect), so that a disease-free restriction would preferentially remove individuals whose low activity is disease-driven and bias the instrument set in an unpredictable direction. Including all eligible participants regardless of morbidity therefore preserves the representativeness of the exposure instruments and guards against this form of selection bias, at the cost of a more phenotypically heterogeneous exposure sample. Participant characteristics are in Supplementary Table 1.

#### Outcome (molecular layers)

For each molecular layer, summary statistics of the SNPs significantly associated with VPA were extracted. Summary statistics included SNPs, beta effect sizes, and p values. Sources are outlined below:

**Table 1.** Sources of omics data used as outcomes in the multi-omic MR-GAT framework

| Layer | Tissue | Dataset | Sample size | Ancestry |
| --- | --- | --- | --- | --- |
| Proteomic | Plasma | UKBB Pharma Proteomics Project | ~54,000 | Predominantly European |
| Epigenomic (CpG methylation) | Whole blood | GoDMC | ~27,750 | European |
| Glycomic | Plasma | TPNG GWAMA (plasma N-glycome) | ~10,000 | European |
| Bulk transcriptomic | Whole blood | eQTLGen | ~31,684 | European |
| Single-cell transcriptomic | PBMCs | OneK1K | 982 | European |

#### Network

Gene-gene interactions were defined using the STRING protein-protein interaction database (version 12.0; Szklarczyk et al., 2023), retaining high-confidence interactions (combined score ≥700).

#### Reference gene sets (post-hoc interpretation)

Two independently derived gene sets were used to interpret the prioritised genes: ageing-causal genes from the epigenome-wide MR analysis of Ying et al. (2024), and exercise-responsive genes from the MoTrPAC human study (MoTrPAC Study Group, 2026).

The MoTrPAC acute endurance bout comprised 40 minutes of continuous cycling at ∼65% VO₂peak (an intensity characterised as ‘vigorous’ for the sedentary cohort studied; n = 64 participants completed the baseline acute endurance bout), whereas the UKBB VPA phenotype (≥6 METs, also characterised as ‘vigorous’; Cai et al., 2026; Garber et al., 2011) represents habitual, chronic activity. MoTrPAC is accordingly used here only as an external index of whether the prioritised genes are exercise-responsive in general; the overlap should be read as evidence of broad exercise-responsiveness rather than as validation of the chronic, habitual activity response. Genes and proteins significantly responsive (adjusted p < 0.05) in the acute exercise-versus-control contrast were extracted from the MoTrPAC blood transcriptome and Olink proteome differential analysis results (5,040 genes and proteins in total; 948 within the tested universe; MoTrPAC Study Group, 2026).

The acute data was used in preference to MoTrPAC’s chronic training data for three reasons: the chronic molecular (transcriptomic and proteomic) differential analysis datasets from this cohort were not yet released at the time of analysis (only the acute contrasts were available); the corresponding chronic physiological cohort was small and truncated by the COVID-19 suspension (only 45 of 206 randomised participants completed post-intervention follow-up testing, with as few as ∼14-18 per exercise arm), rendering any chronic molecular contrast severely underpowered; and the chronic MoTrPAC contrast reflects trained versus untrained adaptation to a structured 12-week programme, which would not cleanly correspond to the germline-anchored liability to habitual activity that the UKBB exposure captures.

Genes causally implicated in biological ageing were obtained from Ying et al. (2024), who performed epigenome-wide Mendelian randomisation (EWMR) across 420,509 CpG sites, using whole-blood methylation quantitative trait loci (GoDMC; 27,750 individuals) as instruments and eight lifespan and healthspan-related traits as outcomes, retaining CpGs with FDR-significant causal effects supported by colocalisation. These causal CpGs are distinct from the sites prioritised by conventional age-correlative epigenetic predictors (e.g. Horvath, Hannum, PhenoAge, GrimAge); indeed, Ying et al. showed that these conventional predictors are not enriched for the causal CpGs. The ageing-causal reference set used here was constructed from the subset of these causality-enriched CpGs that retained FDR-significant, colocalised causal associations in the EWMR results, mapped to genes via the Illumina UCSC RefGene annotation, yielding 183 genes (33 within the tested universe). Because this reference is derived from germline-instrumented, pleiotropy-filtered MR rather than from correlation with chronological age, its comparison with the VPA-anchored genes constitutes a like-for-like convergence of two independent causal analyses, and not an overlap with a correlative ageing clock.

### Exome-Wide Association Study (ExWAS)

An exome-wide single-variant association study was performed on the UKBB Research Analysis Platform using OQFE WES data to identify protein-coding variants associated with habitual vigorous physical activity (VPA).

#### Phenotype

VPA was defined using the UKBB-derived accelerometry measure (Field 90139) as the fraction of time spent in VPA (>400 milligravities corresponding to ≥6 METs; Cai et al., 2026), measured over a seven-day wrist-worn accelerometer wear period in participants with valid wear time, adequate calibration, and uninterrupted recording. For the ExWAS, VPA was designated as the outcome and analysed adjusted for sedentary activity, which was included as a covariate.

#### Sample quality control

Analyses were restricted to participants of European (Caucasian) genetic ancestry. Of approximately 93,000 participants with valid accelerometry, individuals were excluded for unusually high heterozygosity or genotype missingness >5% (n = 131), sex-chromosome aneuploidy (n = 57), mismatch between self-reported and genetically inferred sex, and relatedness to other participants. Approximately 78,000 participants remained after quality control, of whom approximately 75,000 had WES data and constituted the analysis sample. Genotype data were lifted over from GRCh37 to GRCh38, and low-quality variants in the SNP-array and WES data were removed using default filters.

#### Covariates

Association testing used a linear regression model (REGENIE, fitted for the quantitative VPA trait). In addition to the sedentary activity adjuster, the model accounted for age, sex, UK Biobank assessment centre, season of accelerometer wear, body mass index (BMI), and Townsend Deprivation Index.

#### Association testing

Single-variant association testing was conducted using REGENIE. In Step 1, a whole-genome ridge regression model was fitted using the genome-wide SNP-array genotypes to capture polygenic background and population structure; in Step 2, single-variant association tests of the WES variants for the quantitative VPA phenotype were performed conditional on the Step 1 leave-one-chromosome-out predictions, fitting linear regression across the autosomal chromosomes (1–22). The conventional genome-wide significance threshold (p < 5 × 10⁻□) was applied.

#### Fine mapping and instrument selection

Genome-wide significant loci were processed in LocusZoom, and regional posterior probabilities of association were used to guide selection of the lead instruments, yielding 86 SNPs carried forward into the downstream MR. Correlated instruments within a locus were subsequently handled explicitly through the LD-aware MR framework below.

### LD-Aware, Overlap-Aware Mendelian Randomisation

Causal effects of VPA on each gene were estimated using two-sample MR, in which the validity of causal inference rests on three core assumptions: that the genetic instruments are robustly associated with the exposure (relevance); that they share no common cause with the outcome (independence); and that they influence the outcome only through the exposure (exclusion restriction). The analytical procedures below were designed to satisfy, and where possible to test, each of these assumptions in turn.

#### Instrument relevance

The exposure instruments were the VPA-associated variants identified in the ExWAS at genome-wide significance (p < 5 × 10⁻□), ensuring a robust statistical association between each instrument and the exposure. To avoid weak instrument bias, analyses were restricted to genes with at least two overlapping, LD-referenced instruments, and instruments were drawn only from the fine mapped lead variant set, prioritising variants with strong and credible exposure associations.

#### Independence and the handling of linkage disequilibrium

Because correlated instruments at the same locus can bias the causal estimate and violate the assumption of independent instrument contributions, LD was modelled explicitly rather than assumed away. For each omic layer, exposure variants were harmonised to the outcome summary statistics by intersecting SNP identifiers, aligning alleles, and flipping outcome effect estimates where alleles were reversed, ensuring that exposure and outcome effects referred to the same effect allele. Harmonised variants were then intersected with the LD reference panel to ensure availability in the LD matrix. Causal estimates were computed using an LD-aware inverse variance weighted (IVW) estimator. Given vectors of exposure effects and outcome effects across SNPs and the corresponding LD matrix, a covariance matrix was defined as:

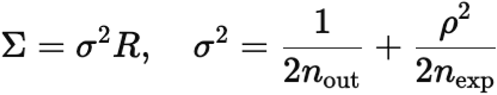

where is the exposure sample size, is the outcome sample size, and is a cross-trait correlation parameter. The LD-aware IVW estimate was then:

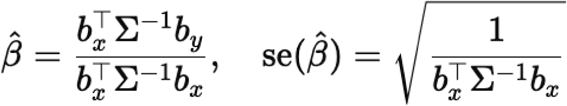

with Wald and two-sided -values. Overlap-aware correction was applied only to UKBB outcomes (proteomic layer); all other datasets were analysed using an LD-aware IVW framework assuming no sample overlap.

#### Exclusion restriction and the control of horizontal pleiotropy

The exclusion restriction assumption (that instruments affect the outcome only through the exposure) is violated by horizontal pleiotropy, in which a variant influences the outcome through pathways independent of VPA. This was tested directly using the MR-Egger intercept, which provides a formal test of directional (unbalanced) pleiotropy. Gene-level estimates showing evidence of directional pleiotropy (MR-Egger intercept p < 0.05) were excluded, so that only instruments with no detectable directional pleiotropy were retained. This pleiotropy filtering procedure was applied uniformly across all molecular layers, ensuring that the genes carried forward to the multi-omic trait importance score satisfied the exclusion restriction assumption as far as it is testable.

- -

-

_X_] (an intercept column and the SNP-exposure effects), and the LD-aware weight matrixS^-1^¹ = (D Ω D)^-1^ (where D is the diagonal of outcome standard errors and Ω the LD matrix):

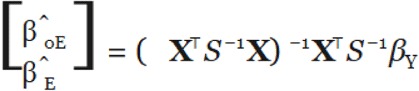

0E -

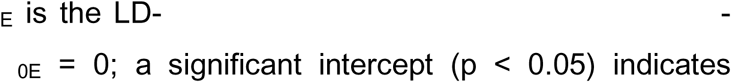

#### Residual limitations

As is standard in MR, only the relevance assumption is fully verifiable; independence and exclusion restriction can be supported and tested but not proven. The MR-Egger intercept tests directional pleiotropy but cannot exclude balanced pleiotropy, and the LD-aware and overlap-aware corrections mitigate but do not wholly eliminate bias from correlated instruments or sample overlap. These estimates are therefore interpreted as causally-anchored rather than definitively causal.

### Multi-Omic Trait Importance (MTI) Score

Having estimated gene-level causal effects of VPA across the five molecular layers by MR, these per-layer MR effect sizes were aggregated into a single continuous score, the multi-omic trait importance (MTI), which served as the supervised training target for the graph model. The MTI was constructed to preserve graded evidence across layers and to avoid artefacts introduced by nominal significance thresholding.

Within each omic layer, MR effect sizes were z-scored across genes to ensure comparable scaling between molecular modalities. The continuous MTI score for each gene was then computed as:

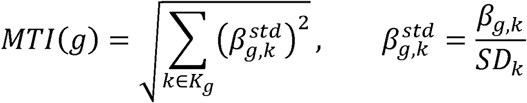

where β_g,k_ denotes the MR effect size for gene *g* in omic layer *K*, SD_K_ is the standard deviation of effect sizes within layer *k* and *k*_g_ is the set of omic layers with available MR evidence for that gene. All layers were weighted equally; no layer-specific weighting was applied. Because the normalised effect sizes are squared before summation, the MTI is non-negative and reflects the magnitude of multi-layer causal association irrespective of the direction of effect, prioritising genes with strong causal signal in any direction across multiple layers.

For sensitivity analyses, alternative MTI variants were computed using nominal (p < 0.05) and false discovery rate (FDR < 0.05) thresholds; however, all primary analyses and downstream gene prioritisation used the continuous MTI formulation. The MTI thus represents the per-gene regression target that the graph model is trained to reconstruct from network context, rather than an independent result in itself.

### Graph Construction

The prioritisation framework was structured as a gene interaction graph, in which each node represented a gene and each edge a protein-protein interaction, allowing the model to integrate per-gene causal evidence with the topology of the wider interaction network.

#### Nodes

Each gene was represented as a node whose feature vector encoded its multi-omic causal evidence: the standardised gene-level MR effect sizes from each molecular layer. The multi-omic trait importance (MTI) score and its derived quantities were not included as node features but served as the supervised regression target; a leakage guard explicitly excluded all MTI-derived columns from the feature matrix to prevent target leakage. Features were z-scored across genes, and where a gene lacked MR evidence in a given layer, the corresponding value was set to zero.

The chromosome 17q21.31 inversion region was excluded from the node set prior to graph construction, owing to its extended, atypical linkage disequilibrium structure and high pleiotropy, which can distort both instrument selection and network connectivity; all reported node and universe counts are post-exclusion.

#### Edges

Gene-gene edges were defined using the STRING protein-protein interaction database (version 12.0; Szklarczyk et al., 2023), retaining only high-confidence interactions with a combined confidence score ≥700. Edges were treated as unweighted, with the confidence threshold applied as a binary inclusion criterion.

Genes lacking any high-confidence interaction were retained as nodes but contributed no edges; the graph attention network propagates over edges while still scoring isolated nodes from their features. The connected component comprised 2,473 genes joined by 17,193 high-confidence edges, and this connected STRING graph constituted the tested gene universe for all GAT rank-based enrichment analyses. The chromosome 17q21.31 region was excluded during node construction (see Nodes), so neither the graph nor the downstream universes include genes from this locus.

### GAT Architecture, Training, and Ranking

A two-layer graph-attention network (GAT) encoder was used, consisting of a graph-attention convolution layer (GATConv), an exponential linear unit (ELU) activation, dropout regularisation, and a second graph attention convolution layer (GATConv → ELU → Dropout → GATConv), followed by two heads sharing the same node embeddings: (i) a regression head (a linear layer) predicting the continuous multi-omic trait importance (MTI) score; and (ii) a classification head (a linear layer) predicting a logit for multi-layer support. The multi-layer support label was defined as:

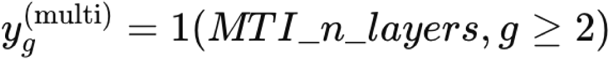

The total loss was:

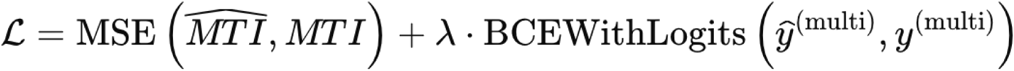

with a class-imbalance pos_weight applied to the BCE term (capped at 50) and

. The model was trained with Adam (lr) for 300 epochs.

After training, MTI-only ranking was generated by sorting in descending order and computing for a hybrid score combining z-scored predictions:

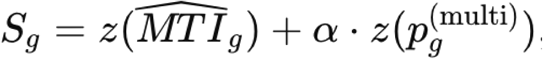

where and is user-specified (default 1.0 in the training script). Genes were ranked by in descending order to produce the final hybrid GAT rank.

### Overlap with Exercise-Responsive and Ageing-Causal Genes

Two distinct background universes are defined and kept separate throughout. The first is the connected STRING graph (N = 2,473 genes, each carrying at least one high-confidence edge), which is the universe for all GAT rank-based enrichment, since only connected genes receive graph-propagated rankings. The second is the multi-omic MR universe (N = 2,959 genes possessing a multi-omic trait-# importance score across the five molecular layers), which is the universe for the model-free convergence between the two causal gene sets; this test is not graph-based and is therefore not restricted to connected nodes. The two universes differ because a gene may carry multi-omic MR evidence (and thus an MTI score) without having a high-confidence STRING interaction; the connected graph is a subset of the MTI-scored genes. Each reference set was intersected with the relevant universe before testing, so that the drawn sample and the reference set were always defined within the same background.

To situate the GAT-prioritised genes within exercise and ageing biology, their ranking was tested for statistical enrichment against two independently derived reference gene sets. These analyses were conducted strictly as post-hoc interpretative assessments and were not used for model training, optimisation, or supervision.

Genes responsive to acute exercise were obtained from the MoTrPAC human study (MoTrPAC Study Group, 2026), using the whole-blood differential-analysis results. Genes showing significant differential abundance (adjusted p < 0.05) in the acute exercise versus control contrast were extracted from the blood transcriptomic and blood Olink proteomic layers and combined into a single exercise-responsive set. As the MoTrPAC contrast reflects the acute response to a single moderate-to-vigorous exercise bout rather than chronic adaptation to vigorous activity (as in UKBB), this set represents exercise-responsiveness in general and is not interpreted as a measure of chronic adaptation. Genes causally implicated in biological ageing were obtained from the supplementary files of Ying et al. (2024).

### Background and gene universe

GAT rank-based enrichment tests were evaluated against the connected STRING graph (N = 2,473 genes); the model-free convergence test was evaluated against the multi-omic MR universe (N = 2,959 MTI-scored genes). In each case the reference set was intersected with the relevant universe before testing, rather than against the whole genome, to avoid inflation arising from differences in gene-set composition.

#### Enrichment testing

For each reference set, enrichment among the top-ranked GAT genes was assessed using the hypergeometric test (equivalently, a one-sided Fisher’s exact test). Let *N* be the number of genes in the tested universe, *M* the number of reference-set genes present in that universe, *K* the number of top-ranked genes drawn from the ranking, and *x* the number of reference-set genes observed among those top *K*. Under the null hypothesis of no association between ranking and reference-set membership, the probability of observing exactly *i* reference genes among the top *K* is given by the hypergeometric distribution:

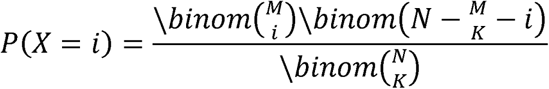

The enrichment *p*-value is the one-sided upper tail probability of observing at least as many reference genes as were seen:

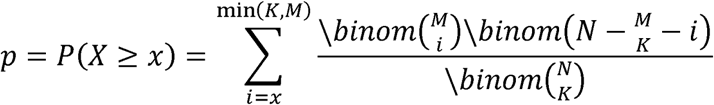

The number of reference genes expected among the top K under the null is:

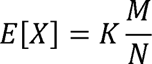

and the fold-enrichment reported throughout is the ratio of observed to expected counts:

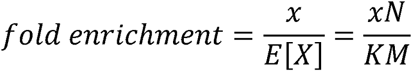

To assess robustness, enrichment was computed across a range of rank thresholds (K = 100, 150, 200) and across the five random model initialisations, with a signal considered robust if significant in the majority of seeds. The identical procedure was applied to alternative ranking strategies (raw MR p-value ranking, MR effect-size ranking, MTI ranking, and a multilayer-perceptron or MLP baseline) to determine whether any enrichment recovered by the GAT was also recoverable without graph-based propagation. Enrichment p-values were therefore interpreted relative to these baselines.

#### Model-free convergence

In addition to the ranking-based enrichment, a model-free convergence test was performed directly between the two causal sets, using the same hypergeometric formulation but with the drawn set defined by causal significance rather than by model rank. Here the “sample” is the set of FDR-significant exercise-MR genes (FDR < 0.05; *n* = 906), the reference set is the ageing-causal genes present in the universe (M = 33), and x is their observed overlap, evaluated against the hypergeometric null with the multi-omic MR universe (N = 2,959 MTI-scored genes) as background. The expected overlap under the null is E[X] = n · M / N = 906 × 33 / 2,959 ≈ 10.1, against the 16 observed (1.6-fold; p = 0.023).

### Two-sample Mendelian randomisation across four ageing outcomes

#### Outcome datasets

We selected four genetically distinct ageing-related outcomes spanning the lifespan-healthspan axis, mirroring the trait panel used in prior epigenome-wide causal ageing studies (Ying et al., 2024). Parental lifespan was taken from Timmers et al. (2019), a genome-wide association study of the combined survival of both parents, with effects in units of negative log-hazard. Healthspan was taken from Zenin et al. (2019; N = 300,447 UK Biobank participants; age at first incidence of a major age-related morbidity). The first genetic principal component of ageing (aging-GIP1) which is a composite of healthspan, parental lifespan, exceptional longevity, frailty and self-rated health, and the trait most strongly genetically correlated with healthy lifespan, was taken from Timmers et al. (2022), with effects in standardised units. Exceptional longevity was taken from Deelen et al. (2019; survival beyond the 90th survival percentile; 11,262 cases versus 25,483 controls), analysed on the case control (log-odds) scale, with case proportion s = 11,262 / 36,745 = 0.31. All four datasets were on genome build GRCh37; lifespan, healthspan and aging-GIP1 are quantitative traits, whereas longevity is binary and was treated as a case control trait in colocalisation. Effect estimates are reported on each outcome’s native scale (hazard, standardised, or log-odds) and are therefore not directly comparable in magnitude across outcomes.

#### Instruments

Each gene was instrumented by its cis-acting regulatory variants. Protein-level instruments were drawn from the UK Biobank Pharma Proteomics Project cis-pQTL (UKB-PPP; full cohort N = 54,219, with CTSF assayed in N = 33,822), available for six of the eight genes (*CTSD, CTSF, HEXIM1, IGFBP7, LTBP3, RHOC*); *FADS1* and *FADS2* were not assayed on the Olink panel. The cis-pQTL coordinates were converted from GRCh38 to GRCh37 by UCSC liftOver prior to harmonisation. Expression-level instruments were drawn from eQTLGen whole-blood cis-eQTL (N ≈ 31,684) for all eight genes.

#### Instrument selection and harmonisation

Within each gene’s cis region, expression instruments (eQTLGen) were LD-clumped (r² < 0.001, 10 Mb) against the 1000 Genomes EUR panel; protein instruments (UKB-PPP), which lack rsIDs, were represented by the genome-wide lead cis-pQTL variant and analysed by the Wald ratio, with cross-layer support assessed by colocalisation from the full regional data. Outcomes were harmonised on chromosome:position and alleles.

#### Causal estimation

For gene-outcome pairs with two or more independent instruments, causal effects were estimated by inverse variance weighted (IVW) regression. For instruments indexed by j, with exposure effects βX,j, outcome effects βY,j, outcome standard errors σY,j, and weights w□ = 1 / σY,j²:

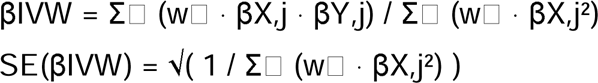

Between-instrument heterogeneity, which can indicate pleiotropy, was quantified by Cochran’s Q, distributed as χ² with k − 1 degrees of freedom for k instruments: Q = Σ□w□· (βY,j − βIVW · βX,j)²

For pairs with a single independent cis instrument, the causal effect was estimated by the Wald ratio, β = βY / βX, with delta method standard error SE = |σY / βX|. Because the single-instrument Wald z-statistic reduces to βY / σY of the lead variant (determined by the outcome association, the exposure effect cancelling), single-instrument MR significance is not independent across instrument layers that share a lead variant. For such pairs, cross-layer support was assessed through colocalisation, computed from the full regional data in each arm, rather than through agreement of Wald p-values. The direction of causality was tested for every signal using the MR Steiger test, retaining only signals in which the instrument explained more variance in the exposure than in the outcome.

For the eQTL arm, instrument effect sizes were obtained by converting eQTLGen Z-scores to β and SE using the minor allele frequency (f) and per-SNP sample size (N):

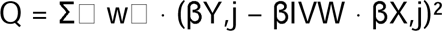

with f taken from the minor allele frequency of the matched outcome variant.

Protein arm estimates used the lead variant Wald ratio throughout; expression arm estimates used IVW where ≥2 LD-clumped instruments were available and the Wald ratio otherwise.

#### Colocalisation

Each gene-outcome pair was tested for colocalisation using the approximate Bayes factor method implemented in coloc (v5.2.3; Giambartolomei et al., 2014), computed from the full cis-regional summary statistics. For a region containing a molecular exposure and an ageing outcome, coloc evaluates five mutually exclusive hypotheses: H₀, no causal variant for either trait; H₁ and H₂, a causal variant for the exposure or the outcome alone, respectively; H₃, distinct causal variants for the two traits; and H₄, a single shared causal variant. Assuming at most one causal variant per trait in the region, a Wakefield approximate Bayes factor was computed for each variant from its effect estimate and standard error and combined with prior probabilities (coloc defaults: p₁ = p₂ = 1 × 10□ that a variant is causal for one trait, p₁₂ = 1 × 10□ that it is causal for both) to yield the posterior probability of each hypothesis (PP.H0–PP.H4). The posterior probability of a shared causal variant, PP.H4, is the quantity of interest for colocalisation.

Quantitative outcomes were analysed as type “quant” and exceptional longevity as a case control trait (type “cc”, case proportion s = 11,262/36,745 = 0.31). Sample sizes supplied to coloc were the study totals for the molecular exposures (cis-pQTL, UKB-PPP N = 33,822; cis-eQTL, eQTLGen N ≈ 31,684) and, for the longevity outcome, the per-variant effective sample size reported in the GWAS (mean ≈ 11,473 across the CTSF cis region); the three quantitative ageing outcomes used their published sample sizes (healthspan N = 300,447; aging-GIP1 and parental lifespan as reported by Timmers et al., 2022 and 2019).

For the expression arm, eQTLGen Z-scores were converted to effect sizes using the minor-allele frequency of the matched outcome variant. Because ageing GWAS are statistically underpowered relative to molecular QTL studies (which inflates the posterior mass on H₀, H₁ and H₂ and weakens the discrimination between H₃ and H₄) we additionally reported the conditional colocalisation probability, conditional PP.H4 = PP.H4 / (PP.H3 + PP.H4), which expresses the probability of a shared causal variant conditional on each trait having a causal variant in the region and is therefore interpretable independently of the outcome GWAS’s power. This conditional formulation follows the principle formalised by Wallace (2020) that colocalisation evidence is more robustly assessed by conditioning on the presence of a causal variant in each trait than by the unconditional posterior alone, and was applied to ageing outcomes as in Ying et al. (2024). Values > 0.7 were taken as evidence of colocalisation (Ying et al., 2024; Timmers et al., 2022).

#### Significance and validation criteria

P-values were corrected across the full gene × outcome matrix within each instrument arm using the Benjamini-Hochberg false discovery rate, with FDR < 0.05 considered statistically significant. A gene–outcome pair was classified as validated only if it satisfied all three criteria: FDR-significant MR, a Steiger-consistent direction, and conditional PP.H4 > 0.7. All analyses were performed in R using the coloc and data.table packages.

## Code availability

All analysis scripts, gene lists, and links to publicly available datasets are at https://github.com/cgjuan01/Causal-deep-learning-GNN-multi-omic.

## Data Availability

All scripts and data used are available online at https://github.com/cgjuan01/Causal-deep-learning-GNN-multi-omic

---

**Supplementary Table 1.** MR Steiger directionality test for FDR-significant gene-outcome pairs.

| Gene | Outcome | Arm | k | $r^2_{\text{exposure}}$ | $r^2_{\text{outcome}}$ | Steiger-consistent |
| --- | --- | --- | --- | --- | --- | --- |
| CTSF | Exceptional longevity | pQTL | 1 | 0.0192 | $2.7 \times 10^{-4}$ | Yes |
| CTSF | Exceptional longevity | eQTL | 6 | 0.0106 | $5.5 \times 10^{-4}$ | Yes |
| CTSF | aging-GIP1 | pQTL | 1 | 0.0192 | $3.3 \times 10^{-4}$ | Yes |
| CTSF | aging-GIP1 | eQTL | 6 | 0.0106 | $2.4 \times 10^{-4}$ | Yes |
| FADS1 | aging-GIP1 | eQTL | 4 | 0.0126 | $2.6 \times 10^{-4}$ | Yes |
| LTBP3 | aging-GIP1 | eQTL | 3 | 0.0152 | $2.8 \times 10^{-4}$ | Yes |
| HEXIM1 | aging-GIP1 | eQTL | 1 | 0.0025 | $1.3 \times 10^{-4}$ | Yes |

*MR Steiger directionality test for all FDR-significant gene-outcome pairs. $r^2_{\text{exposure}}$ and $r^2_{\text{outcome}}$ are the mean variance explained by the cis instruments in the molecular exposure and the ageing outcome respectively; a pair is Steiger-consistent when the instruments explain more variance in the exposure than the outcome. All FDR-significant pairs were Steiger-consistent, so colocalisation was the discriminating validation criterion. eQTL instruments were LD-clumped ( $r^2 < 0.001$ , 10 Mb, 1000G EUR); the protein arm used the lead cis-pQTL variant (rs4930384). Exposure N: eQTLGen 31,684, UKB-PPP 33,822; outcome N: longevity 36,745, aging-GIP1 [insert the printed value].*

## References

1. Amar D, Gay NR, Jimenez-Morales D et al. The mitochondrial multi-omic response to exercise training across rat tissues. Cell Metab. 2024 Jun 4;36(6):1411–1429.e10. doi: 10.1016/j.cmet.2023.12.021.

2. Biswas RK, Ahmadi MN, Bauman A et al. Publisher Correction: Wearable device-based health equivalence of different physical activity intensities against mortality, cardiometabolic disease, and cancer. Nat Commun. 2025 Oct 30;16(1):9581. doi: 10.1038/s41467-025-65754-4.

3. Deelen J, Evans DS, Arking DE et al. A meta-analysis of genome-wide association studies identifies multiple longevity genes. Nat Commun. 2019 Aug 14;10(1):3669. doi: 10.1038/s41467-019-11558-2. Erratum in: Nat Commun. 2021 Apr 23;12(1):2463. doi: 10.1038/s41467-021-22613-2.

4. Doherty A, Smith-Byrne K, Ferreira T et al. GWAS identifies 14 loci for device-measured physical activity and sleep duration. Nat Commun. 2018;9:5257. doi:10.1038/s41467-018-07743-4.

5. Giambartolomei C, Vukcevic D, Schadt EE et al. Bayesian test for colocalisation between pairs of genetic association studies using summary statistics. PLoS Genet. 2014 May 15;10(5):e1004383. doi: 10.1371/journal.pgen.1004383.

6. Grimm S, Ernst L, Grötzinger N et al. Cathepsin D is one of the major enzymes involved in intracellular degradation of AGE-modified proteins. Free Radic Res. 2010 Sep;44(9):1013–26. doi: 10.3109/10715762.2010.495127.

7. Heasman SJ, Ridley AJ. Mammalian Rho GTPases: new insights into their functions from in vivo studies. Nat Rev Mol Cell Biol. 2008 Sep;9(9):690–701. doi: 10.1038/nrm2476.

8. Höhn A, Grune T. Lipofuscin: formation, effects and role of macroautophagy. Redox Biol. 2013 Jan 19;1(1):140–4. doi: 10.1016/j.redox.2013.01.006.

9. Huang J, McLean GR, Franke A. Twenty years of genome-wide association studies: Health translation challenges and AI opportunities. Eur J Hum Genet. 2025;33:1579–1584. doi:10.1038/s41431-025-01951-5.

10. Klimentidis YC, Raichlen DA, Bea J et al. Genome-wide association study of habitual physical activity in over 377,000 UK Biobank participants identifies multiple variants including CADM2 and APOE. Int J Obes (Lond). 2018 Jun;42(6):1161–1176. doi: 10.1038/s41366-018-0120-3.

11. Li X, Liu C, Li W et al. Multi-omics delineate growth factor network underlying exercise effects in an Alzheimer’s mouse model. Alzheimers Dement. 2025 Mar;21(3):e70024. doi: 10.1002/alz.70024. PMID: 40156268;

12. López-Otín C, Blasco MA, Partridge L, Serrano M, Kroemer G. Hallmarks of aging: An expanding universe. Cell. 2023 Jan 19;186(2):243–278. doi: 10.1016/j.cell.2022.11.00

13. Michels AA, Bensaude O. Hexim1, an RNA-controlled protein hub. Transcription. 2018;9(4):262–271. doi: 10.1080/21541264.2018.1429836.

14. MoTrPAC Study Group. Temporal dynamics of the multi-omic response to endurance exercise training. Nature. 2024;629:174–183. doi:10.1038/s41586-023-06877-w.

15. MoTrPAC Study Group, Brandt AR, Fleg J, Goodpaster BH et al. Molecular Transducers of Physical Activity Consortium (MoTrPAC): Initial Insights into the Dynamic Human Responses to Exercise [Preprint]. bioRxiv 2026.03.02.705347; 2026 [cited 2026 Jun 16]. Available from: 10.64898/2026.03.02.705347

16. Qi G, Dutta D, Leroux A et al. Genome-wide association studies of 27 accelerometry-derived physical activity measurements identified novel loci and genetic mechanisms. Genet Epidemiol. 2022 Mar;46(2):122–138. doi: 10.1002/gepi.22441.

17. Ristow M, Schmeisser S. Extending life span by increasing oxidative stress. Free Radic Biol Med. 2011 Jul 15;51(2):327–36. doi: 10.1016/j.freeradbiomed.2011.05.010.

18. Robertson IB, Horiguchi M, Zilberberg L, Dabovic B, Hadjiolova K, Rifkin DB. Latent TGF-β-binding proteins. Matrix Biol. 2015 Sep;47:44–53. doi: 10.1016/j.matbio.2015.05.005.

19. Schaeffer L, Gohlke H, Müller M et al. Common genetic variants of the FADS1 FADS2 gene cluster and their reconstructed haplotypes are associated with the fatty acid composition in phospholipids. Hum Mol Genet. 2006 Jun 1;15(11):1745–56. doi: 10.1093/hmg/ddl117

20. Serhan CN. Pro-resolving lipid mediators are leads for resolution physiology. Nature. 2014 Jun 5;510(7503):92–101. doi: 10.1038/nature13479.

21. Shenhar B, Frenkl S, Levy T, Alon U. Maximal human lifespan in light of a mechanistic model of aging. bioRxiv. Published online December 23, 2025. doi:10.64898/2025.12.22.695887.

22. Siintola E, Partanen S, Strömme P, Haapanen A, Haltia M, Maehlen J, Lehesjoki AE, Tyynelä J. Cathepsin D deficiency underlies congenital human neuronal ceroid-lipofuscinosis. Brain. 2006 Jun;129(Pt 6):1438–45. doi: 10.1093/brain/awl107.

23. Singh R, Kaushik S, Wang Y, Xiang Y, Novak I, Komatsu M, Tanaka K, Cuervo AM, Czaja MJ. Autophagy regulates lipid metabolism. Nature. 2009 Apr 30;458(7242):1131–5. doi: 10.1038/nature07976.

24. Smith KR, Dahl HH, Canafoglia L et al. Cathepsin F mutations cause Type B Kufs disease, an adult-onset neuronal ceroid lipofuscinosis. Hum Mol Genet. 2013 Apr 1;22(7):1417–23. doi: 10.1093/hmg/dds558.

25. Spence JP, Mostafavi H, Ota M et al. Specificity, length and luck drive gene rankings in association studies. Nature. 2025 Nov 5. doi: 10.1038/s41586-025-09703-7.

26. Sun BB, Chiou J, Traylor M et al. Plasma proteomic associations with genetics and health in the UK Biobank. Nature. 2023 Oct;622(7982):329–338. doi: 10.1038/s41586-023-06592-6.

27. Sun ED, Zhou OY, Hauptschein M et al. Spatial transcriptomic clocks reveal cell proximity effects in brain ageing. Nature. 2025;638:160–171. doi:10.1038/s41586-024-08334-8.

28. Timmers PR, Mounier N, Lall K et al. Genomics of 1 million parent lifespans implicates novel pathways and common diseases and distinguishes survival chances. Elife. 2019 Jan 15;8:e39856. doi: 10.7554/eLife.39856.

29. Timmers PRHJ, Wilson JF, Joshi PK, Deelen J. Multivariate genomic scan implicates novel loci and haem metabolism in human ageing. Nat Commun. 2020 Jul 16;11(1):3570. doi: 10.1038/s41467-020-17312-3.

30. Vinyard ME, Strobel B, Schiebinger G, et al. Learning cell dynamics with neural differential equations. Nat Mach Intell. 2025;7:1969–1984. doi:10.1038/s42256-025-01150-3.

31. Võsa U, Claringbould A, Westra HJ et al. Large-scale cis- and trans-eQTL analyses identify thousands of genetic loci and polygenic scores that regulate blood gene expression. Nat Genet. 2021 Sep;53(9):1300–1310. doi: 10.1038/s41588-021-00913-z.

32. Yang H, Roy G, Nguyen AQ, et al. MultiCell: geometric learning in multicellular development. Nat Methods. 2025. doi:10.1038/s41592-025-02983-x.

33. Ying K, Liu H, Tarkhov AE, Sadler MC, Lu AT, Moqri M, Horvath S, Kutalik Z, Shen X, Gladyshev VN. Causality-enriched epigenetic age uncouples damage and adaptation. Nat Aging. 2024 Feb;4(2):231–246. doi: 10.1038/s43587-023-00557-0.

34. Wajapeyee N, Serra RW, Zhu X, Mahalingam M, Green MR. Oncogenic BRAF induces senescence and apoptosis through pathways mediated by the secreted protein IGFBP7. Cell. 2008 Feb 8;132(3):363–74. doi: 10.1016/j.cell.2007.12.032.

35. Wallace C. Eliciting priors and relaxing the single causal variant assumption in colocalisation analyses. PLoS Genet. 2020 Apr 20;16(4):e1008720. doi: 10.1371/journal.pgen.1008720. Erratum in: PLoS Genet. 2026 May 19;22(5):e1012155. doi: 10.1371/journal.pgen.1012155.

36. Williamson J, Hughes CM, Burke G, Davison GW. A combined γ-H2AX and 53BP1 approach to determine the DNA damage-repair response to exercise in hypoxia. Free Radic Biol Med. 2020 Jul;154:9–17. doi: 10.1016/j.freeradbiomed.2020.04

37. Zenin A, Tsepilov Y, Sharapov S, et al. Identification of 12 genetic loci associated with human healthspan. Commun Biol. 2019;2:41. Available from: 10.1038/s42003-019-0290-0

